# Group Response Analysis: Clinically Interpretable Longitudinal Responder Analysis Methods Developed Using FDA Data

**DOI:** 10.1101/2025.08.13.25333581

**Authors:** Ian DeLorey, Warren B. Bilker, Darina Chudnovskaya, Andrew Conroy, Tara McWilliams, Christopher Miller, Charles E. Argoff, Ian Barnett, Russel Bell, Jennifer Haythornthwaite, Jennifer S. Gewandter, Ian Gilron, Nathaniel P. Katz, Katherine N. Theken, John T. Farrar

**Author notes:** Contact: John T. Farrar, MD, PhD, Professor of Epidemiology, Neurology, and Anesthesia, Room 816 Blockley Hall, University of Pennsylvania, Philadelphia, PA 19104.

## Abstract

Responder analyses for the evaluation of randomized clinical trial (RCT) data have become more common in the recent past, since they can provide the medical community with results that are more directly applicable to clinical care. For pain studies, the predominant responder analysis compares the change in the individual participants’ pain level at baseline to their value at the end of the study period and uses a predetermined clinically important change cut-off value to define a response. While useful, this method substantially reduces the efficiency of the RCT by dichotomizing the results and is limited to comparing the baseline to the end of the study only. In this paper, we introduce a novel approach to the patient response over time with a focus on single dose post-operative studies. This technique provides graphical presentations and statistical approaches to understand the onset of any specified level of response, the maximum proportion of patients with a response at any point in time, and the duration of that response over time. In addition, each outcome can be summarized to examine the result across all possible cut-off points for clinically important differences (CID). We accomplish this by introducing three interrelated, longitudinal efficacy statistics: ROOT, GRO, and GROOT. The response outcome over time (ROOT) estimates the total proportion of a study period an individual patient spends as a responder. The group response outcome (GRO) estimates the instantaneous proportion of responders at all time points across the study period. The group response outcome over time (GROOT) summarizes total efficacy in a cohort, and can be calculated as the area under the GRO curve, or as the mean ROOT; they are identical.

This novel method provides a clinically interpretable responder analysis over the full period of the study and, by using every data point across time, mitigates the loss of statistical power typically associated with dichotomized responder outcomes. Group response analysis is based upon repeated assessments of categorical or continuous measures categorizing each participant’s status as a treatment responder or non-responder at every timepoint based on the prespecified clinically important difference. Both the visual and statistical comparison of any two or more curves provide a comparison of the overall efficacy, which can be statistically tested using a standard asymptotic hypothesis test (such as Wald (Johnson & Romer, 2016)). The method allows for an integrated evaluation of three main components of drug efficacy: the proportion of participants achieving a CID over time (effect), the time to achieve that response (onset), and the length of the response (duration). In this paper, we present the group response analysis methodology and then illustrate it using data from a placebo-controlled randomized clinical trial (RCT) for postoperative pain after third molar extraction treated with meloxicam and ibuprofen as an active comparator (Christensen et al., 2018). Our approach yields similar effect sizes as the sum of pain intensity differences (SPID) commonly used for pain study analyses while providing superior clinical interpretability and a more complete evaluation of drug therapies beyond just efficacy. We propose that this method can be used as a primary or secondary analysis of pain RCTs to answer the question of the patient response to treatment and provide suitable data to compare efficacies across treatment groups.

## 2 Introduction

Originally acute pain clinical trials were analyzed by comparing mean or median values of pain outcomes between randomized groups while the background for responder analyses were also outlined. (Laska et al., 1967; Meier et al., 1958; Sunshine et al., 1964) While initially easier to compute, the group mean values did not easily translate the study results into clinical practice. Given the large variability in participants’ pain experience and response to treatment, a primary goal is to establish the probability that individuals will achieve a clinically important difference (CID) or clinically meaningful response (CMR) to treatment. Over the past 25 years, researchers have increasingly focused on examining individual participant responses and comparing groups as the proportion of participants who achieve a CMR. This shift has led to numerous studies exploring suitable CMR cutoffs for various treatments and diseases. (Dworkin et al., 2009; Farrar et al., 2001; Laigaard et al., 2021; Mathai et al., 2012; Myles et al., 2017; Wyrwich et al., 2004) Often termed a responder analysis, these methods evaluate whether a participant has responded between the beginning and end of a clinical trial. While these approaches have enhanced clinical applicability, the reporting of response over time in clinical trials remains limited.(Mercieca-Bebber et al., 2018; Moore et al., 1996) More recently, this has become an increasingly common analysis in studies of treatments for disease symptoms, particularly acute pain.(Laigaard et al., 2021; Myles et al., 2017) However, responder analyses are often presented as secondary analyses, given their reduced power to detect small differences compared to analyses using continuous outcomes.(Collister et al., 2021)

In this paper, we present a novel methodology developed for the analysis of acute pain studies comprising three interrelated responder outcomes for the analysis of longitudinal, participant-level data. They are: (1) the response outcome over time (ROOT), which estimates the total proportion of study time spent as a responder by an individual; (2) the group response outcome (GRO) at each time point estimates the instantaneous proportion of responders in a cohort; and (3) the GRO over time (GROOT) – calculated identically as either the area under the GRO curve, or as the mean ROOT. The GROOT summary statistic produces similar effect sizes to current analytic approaches while offering several advantages, including a standardized scale for comparative analyses among treatments and the easier clinical interpretability that comes from a responder analyses. They integrate the patient-level components of clinical efficacy (effect, onset, and duration) in a statistically efficient and clinically useful approach. We present an illustrative example using data from a placebo-controlled randomized clinical trial (RCT) for postoperative pain after third molar extraction treated with meloxicam compared to placebo and ibuprofen as an active comparator (Christensen et al., 2018) making use of individual patient data obtained from the US Food & Drug Administration (FDA) Document Archiving, Reporting, and Regulatory Tracking System (DARRTS).

## 3 Methods (For Expanded Description See Supplemental Material)

The following methodology was developed using participant-level data acquired from the FDA, and the study was approved by the University of Pennsylvania Institutional Review Board.

The standard responder analysis examines the difference between the baseline pain level value and another point in time, usually the end point of the study to determine the proportion of participants who achieve a clinically important difference (CID) over that time period. For our analysis, we use that process to estimate the participant’s response status from the baseline through each time point in the study. Individual response may be summarized across a study with the response outcome over time (ROOT), which estimates the total proportion of study time an individual spends as a responder. The group response outcome (GRO) is a function that estimates the instantaneous proportion of participants counted as responders for any time point in the study. The area under the GRO curve, and the mean ROOT, are identical and we define this value as the group response outcome over time (GROOT) ; a summary statistic for total response in a study period.

### 3.1 The Group Response Outcome (GRO)

To calculate the GRO value for each time point, we determine the response status of each participant at each measured time point and then assess the proportion of responders at each protocol prespecified time intervalover the course of the study. Several components of the analysis need to be specified and disclosed as with any data analysis and are outlined in Table 1 for our example analysis. The two that are particularly important are the choice of the CID to achieve a clinically meaningful response (CMR) and the method for handling the interpolation of data across the variable time points logged in the data for each participant’s recording of their outcome measure. Given that the times of measurement are often slightly different or missing for some points in the study for each participant, a method for interpolation between time points is important. Any specific method for this estimation can be used to accommodate for different types of data and are discussed along with other analytic options below.

**Table 1.**
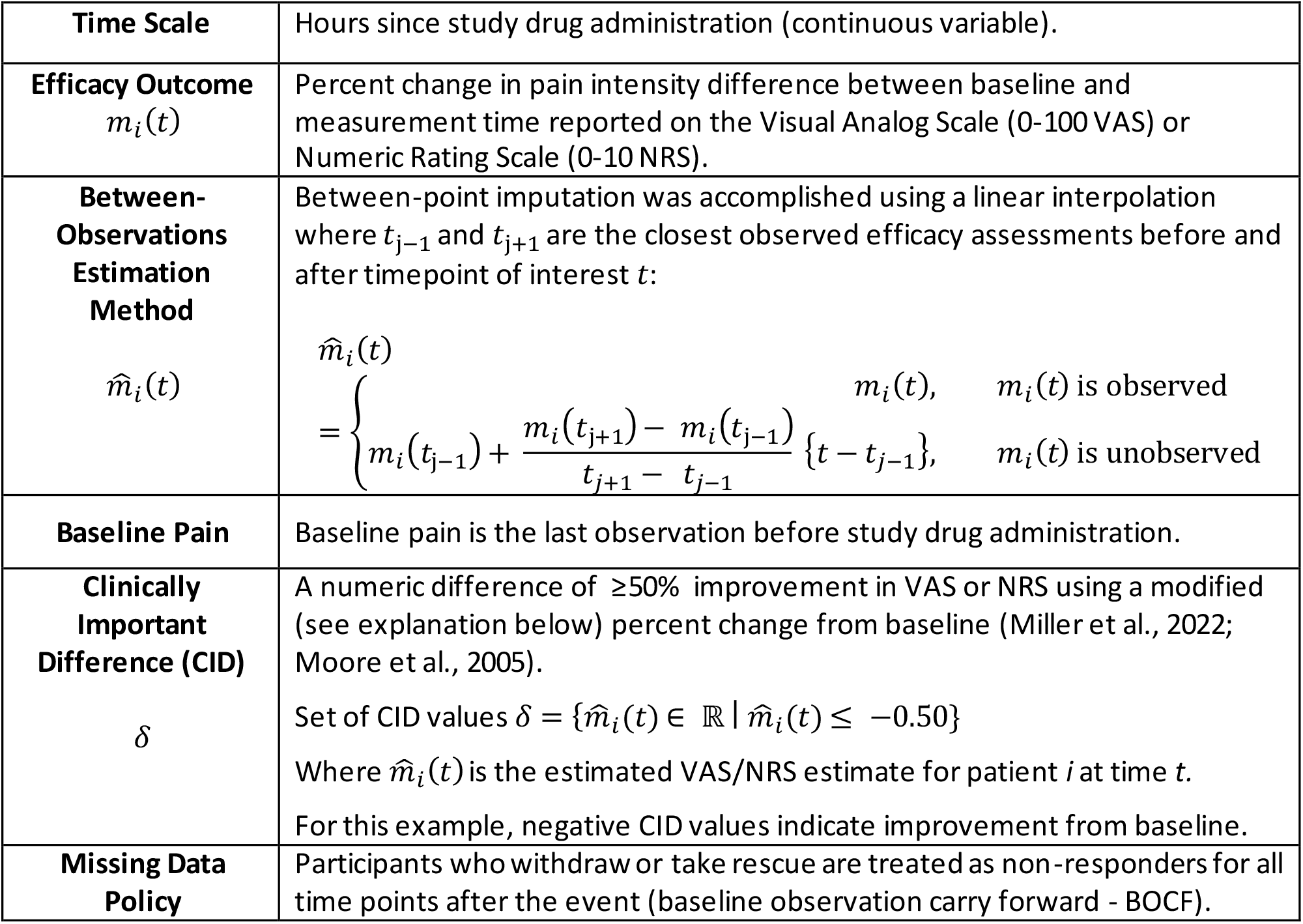
Choices specific to the group response analysis of the example trials.

GRO:

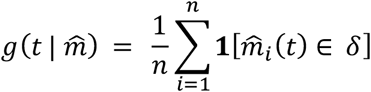

**1**[·] indicator function

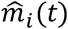 Efficacy outcome estimate for participant *i* at time *t*

*n* number of participants in the group

δ set of clinically meaningful response values or categories

By graphing the proportion of responders over appropriate time intervals, we are able to visualize: (1) onset (rate of rise in the proportion of responders to the maximum number achieving response), (2) maximum proportion of responders (maximum height of the GRO curve) over the full study time, and (3) duration of effect (the proportion of participants who maintain their response over time).

### 3.2 The Response Outcome Over Time (ROOT)

The response outcome over time (ROOT) is the proportion of available time that an individual spends as a responder. For participant *i*, across a study duration that starts at time *a*, and ends at time *b*, and given a function 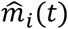 that estimates instantaneous changes in pain from baseline for all times *t* between *a* and *b*:

Root :

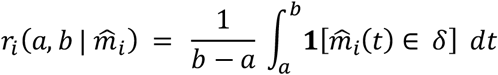

**1**[·] indicator function

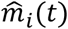 Efficacy outcome estimator for participant *i* at time *t* (see section 3.5)

δ set of clinically meaningful response changes or categories

*a* study start time

*b* study end time

### 3.3 Group Response Outcome over Time (GROOT)

To summarize overall efficacy in a single numeric value, we define the group response outcome over time (GROOT). The GROOT can be calculated as the area under the GRO curve using two methods.

The first is by dividing the value under the GRO curve by the total possible area (study length x 100% relief) of the observed time.

The second is to calculate the proportion of time each participant meets the CMR condition which we define as the response outcome over time (ROOT) and then average those values over all participants.

GROOT:

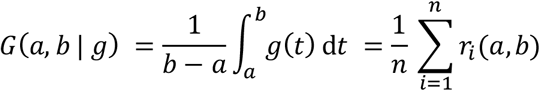

*g*(*t*) group response outcome (GRO) at time *t*

*r*_*i*_(*a, b*) response outcome over time (ROOT) for participant *i*

*a* time interval start

*b* time interval end

Both methods provide identical answers as demonstrated in the supplemental material, but calculation of the confidence interval for the first method requires a simulation process that involves substantial computational effort, while the second can be calculated using familiar asymptotic methods (e.g., Wald or others). The GROOT takes as a value between 0-1, with 1 being 100% of participants obtaining relief for the entire study period. This consistent scale allows a clinically relevant comparison across drug arm dosages. Being a mean value, GROOT is asymptotically normally distributed, but estimation at sample sizes common to acute pain clinical trials may be affected by this restricted range, potentially requiring alternative distributional assumptions.

### 3.4 Calculation of GROOT across all potential CID cut-off points

As with other responder analyses, the GROOT value depends on selecting a CID cutoff value used to define responders. To generalize the GROOT findings and compare the study arms over a range of cutoff values, we can calculate the GROOT and graph the values over all potential cutoff values which can be graphed over the complete range of CID cut-off values for each arm in a study.

### 3.5 Flexible Options for Analysis

One benefit of our novel methodology is the ease with which it may be applied to diverse analytic contexts. Researchers can make selections based on the needs of their own analysis. Table 1 shows choices we made for the application of group response analysis to the FDA acute pain data, with discussion following.

#### 3.5.1 Time Scale and Baseline

GRO estimates the changing level of response over time in the clinical trial, from the defined baseline at time zero, to each time point in the study. It can use any reasonable time scale. For acute pain analgesia, the initial dose of medication administration is an obvious choice for the baseline time zero and the prespecified duration as the final point in the analysis.

#### 3.5.2 Efficacy Outcome and Acceptable CMR Thresholds

These methods require the *a priori* definition of a clinically important difference(CID) to define the clinically meaningful response (CMR). This selection is then applied to individual participant data to dichotomously categorize subjects as a responder or not at each timepoint during the study, using a suitable imputation method to provide a value at any unobserved specific time points. ROOT, GRO & GROOT can be calculated for multiple defined outcome levels and can be graphed across the full range of cut-off values for all treatment groups. (Supplemental Figure A1).

Depending on the specific application, cut-off points can be specified for any outcome, including raw measurements or derived values. Percent change from baseline is common among responder analyses for pain studies(Farrar et al., 2001). Still, researchers must carefully consider the ceiling and floor effect of specific scales and consider using modified percent change outcomes to address the issue that percent changes up and down do not correspond to the same absolute difference.

#### 3.5.3 Missing Data

Participants can withdraw from trials at any time, so the analysis must account for missing data. In this analysis of an acute pain single dose study, we chose to treat patients who took rescue, withdrew, or whose observations otherwise ended early, as non-responders after the event. However, any appropriate missing data method can be adapted. Although it may be that some of those dropping out for side-effects may have experienced successful treatment, the conservative approach is to assume they were non-responders. This prevents bias towards a larger responder rate within study arms. This also reflects the clinical situation where patients who experience significant side effects would likely not want to continue with that treatment in clinical practice even if it was effective.

#### 3.5.4 CMR Status Estimation Between Observations

A GRO that changes only when a participant is actually observed is unlikely to accurately reflect the truth, since the responder status change likely occurs in the period between measurements. For continuous outcomes, a linear imputation of values over time is simple and often appropriate, but any reasonable method may be chosen to estimate these values. Categorical outcomes may require a different estimation method unless it is reasonable to assume a linear relationship between the categories.

### 3.6 Analysis of Onset & Duration

Achievement of CMR can be useful in the analysis of the onset and duration of treatment efficacy. However, simply reporting summary statistics for these values may not provide answers that are clinically relevant. Some participants do not respond to treatment, and for them the concepts of onset and duration cannot be meaningfully applied. One analytical option is to restrict analysis of onset and duration to responders only and is most applicable to the treatment group. Also note that duration can only be meaningfully measured if the study duration is longer than the effect of the treatment. The analysis should be considered observational in nature but answer a relevant clinical question.

Under this framework, onset is summarized as the median time to first achievement of CMR along with the 25%/ 75% range. Duration can be summarized using the time that individuals spend as responders. These definitions are consistent with common clinical questions: (1) “In those patients that achieve a response how long does it take?”; (2) “How much of the treatment period can a responder expect to maintain a response ?”(Supplemental Figure XX). These values provide data to allow clinicians to make an informed decision about the timing of redosing based on the likely time of return of pain in the population.

Observational cohorts based on the post-randomization factor of responder status are not intended to allow a comparison to the placebo group but provide the same type of data gathered in pharmacokinetic studies. In many pain models, it is reasonable to expect a small but significant number of participants to retain CMR for the entire study period. This may constitute informative censoring of duration times, which researchers should consider as they select methods for the analysis of the onset and duration data. A Cure model (Laska et al., 1991) for onset or duration may be a reasonable approach to preserve randomization, handle censoring, and use data from all participants but is beyond the scope of this paper.

## 4 Results

### 4.1 Example Study with IV Meloxicam & Oral Ibuprofen

To illustrate the novel group response analysis methodology, we chose a third molar extraction example study comparing IV meloxicam (15, 30, 60 mg), oral ibuprofen (400 mg) and a placebo, all administered double-dummy. The study was chosen due to its long duration, and variety of treatments.(Christensen et al., 2018)

### 4.2 Visualization of the Group Response Outcome (GRO)

For each study arm included in analysis, we calculate and graph the group response outcome (GRO) over protocol-defined study durations for all treatment arms.

The maximum proportion of concurrent responders (MaxGRO) as shown by the height of the curves, is approximately 80% for the 30 mg and 60mg IV meloxicam group and 68% for the 15mg IV meloxicam and oral ibuprofen groups (Table 2).

**Table 2.**
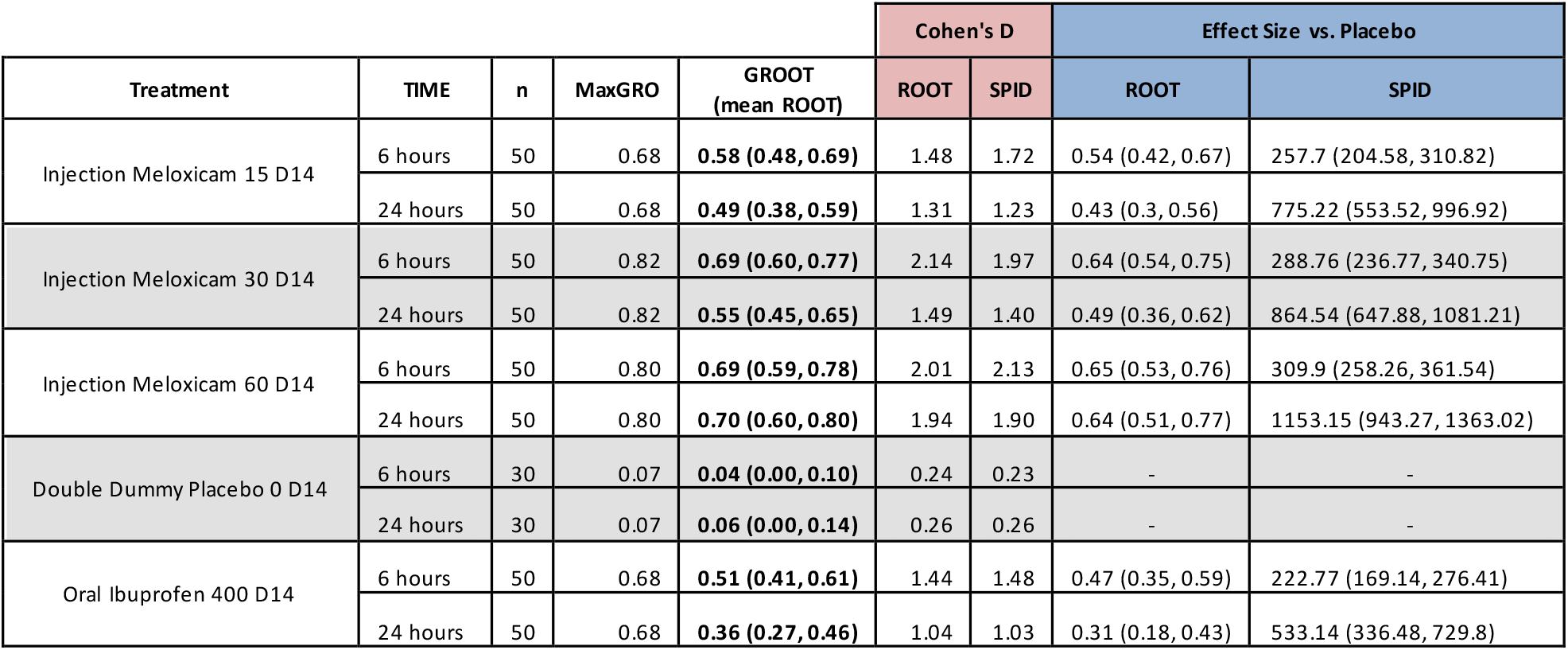
Efficacy summary statistics from group response analysis. GROOT’s standardized scale (0 to 1) allows for easier comparisons than SPID, while producing similar Cohen’s D effect sizes.

| Treatment | TIME | n | MaxGRO | GROOT<br>(mean ROOT) | Cohen's D |  | Effect Size vs. Placebo |  |
| --- | --- | --- | --- | --- | --- | --- | --- | --- |
|  |  |  |  |  | ROOT | SPID | ROOT | SPID |
| Injection Meloxicam 15 D14 | 6 hours | 50 | 0.68 | **0.58 (0.48, 0.69)** | 1.48 | 1.72 | 0.54 (0.42, 0.67) | 257.7 (204.58, 310.82) |
|  | 24 hours | 50 | 0.68 | **0.49 (0.38, 0.59)** | 1.31 | 1.23 | 0.43 (0.3, 0.56) | 775.22 (553.52, 996.92) |
| Injection Meloxicam 30 D14 | 6 hours | 50 | 0.82 | **0.69 (0.60, 0.77)** | 2.14 | 1.97 | 0.64 (0.54, 0.75) | 288.76 (236.77, 340.75) |
|  | 24 hours | 50 | 0.82 | **0.55 (0.45, 0.65)** | 1.49 | 1.40 | 0.49 (0.36, 0.62) | 864.54 (647.88, 1081.21) |
| Injection Meloxicam 60 D14 | 6 hours | 50 | 0.80 | **0.69 (0.59, 0.78)** | 2.01 | 2.13 | 0.65 (0.53, 0.76) | 309.9 (258.26, 361.54) |
|  | 24 hours | 50 | 0.80 | **0.70 (0.60, 0.80)** | 1.94 | 1.90 | 0.64 (0.51, 0.77) | 1153.15 (943.27, 1363.02) |
| Double Dummy Placebo 0 D14 | 6 hours | 30 | 0.07 | **0.04 (0.00, 0.10)** | 0.24 | 0.23 | - | - |
|  | 24 hours | 30 | 0.07 | **0.06 (0.00, 0.14)** | 0.26 | 0.26 | - | - |
| Oral Ibuprofen 400 D14 | 6 hours | 50 | 0.68 | **0.51 (0.41, 0.61)** | 1.44 | 1.48 | 0.47 (0.35, 0.59) | 222.77 (169.14, 276.41) |
|  | 24 hours | 50 | 0.68 | **0.36 (0.27, 0.46)** | 1.04 | 1.03 | 0.31 (0.18, 0.43) | 533.14 (336.48, 729.8) |

### 4.3 GROOT Efficacy Summary Statistic

The group response outcome over time (GROOT) results were calculated as the mean of the participant-level response outcome over time (mean ROOT), showing that the mean proportion of study time individual participants spend with CMR ranges from 0.51 to 0.70 for the range of treatment groups (See Table 2).

All treatment arms are statistically significantly better than placebo (no overlap of the 95% Confidence Interval – 95% CI) for both 6-hour and 24-hour periods. The meloxicam 60mg dose is statistically significantly better than the ibuprofen 400mg dose at 24 hours. For meloxicam 30 mg and 60 mg, the 6-hour GROOT is approximately identical, while the 24-hour GROOT shows separation between the two doses consistent with the differences seen in the GRO curve (Figure 1). The 30mg meloxicam dose has the same time to achieve CMR and MaxGRO value as the 60 mg dose, but the larger dose maintains the benefit for substantially longer, increasing the GROOT. The oral ibuprofen arm has a longer time to achieve CMR of 60 minutes, the same MaxGRO as the 15mg IV meloxicam dose, and a shorter duration,

**Figure 1.**
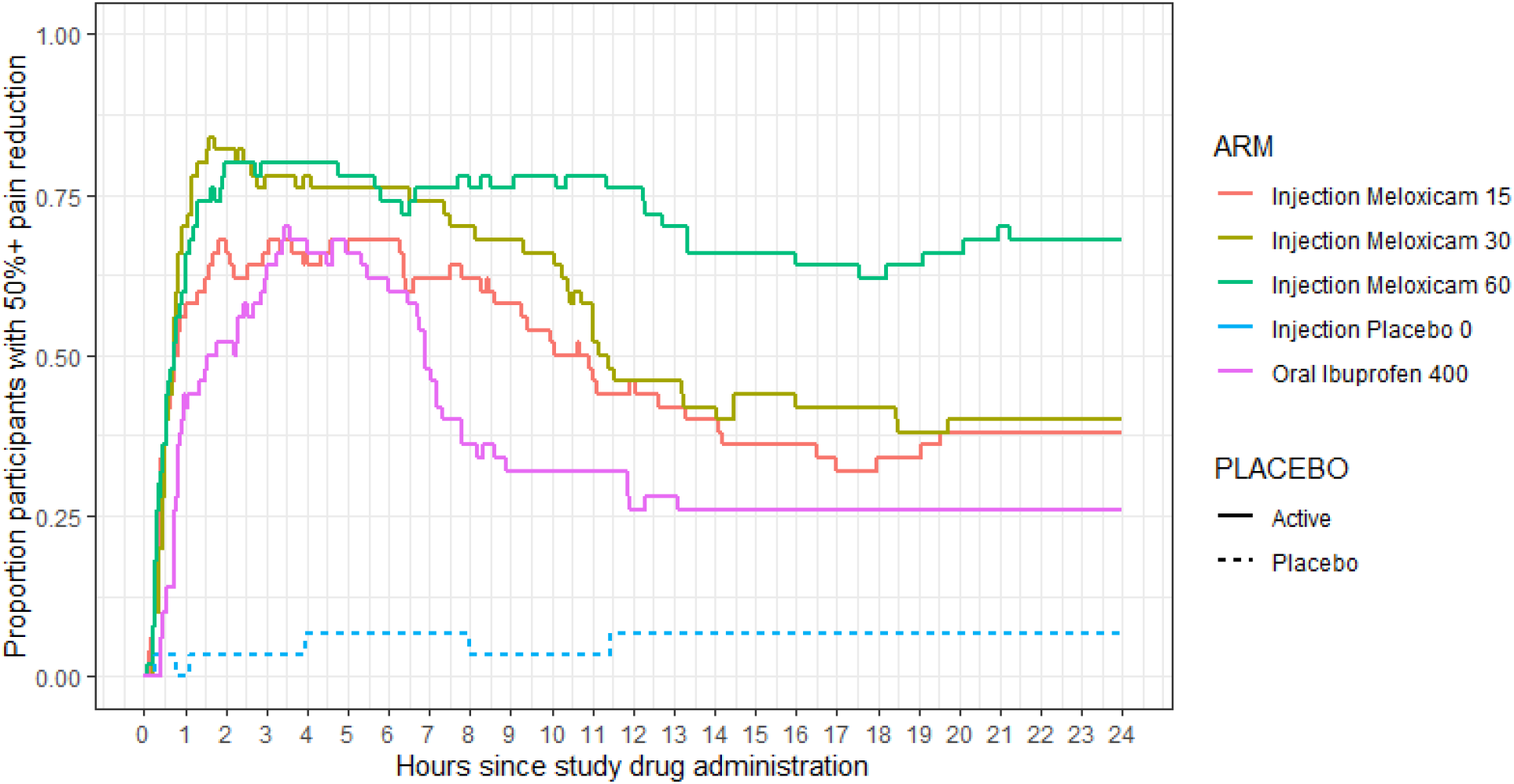
Visualization of the Group Response Outcome (GRO), the changing proportion of participants achieving a ≥50% level (CID) of pain intensity reduction in groups from a single study.

which is consistent with the pharmacokinetic profile of ibuprofen and known duration of its clinical efficacy.

To consider the GROOT findings over a range of cutoff values, we calculated and graphed the values over all potential cutoff levels. (Figure 2). Because this type of graph is limited to a single time point, we selected 6 hours and 24 hours consistent with the drugs’ durations of effect. In the 6-hour graph, the four treatment groups are ordered by dose, and each arm’s GROOT scores decline as the cutoff increases. The ibuprofen arm is slightly lower than all three IV meloxicam doses but provides a reasonable response.

**Figure 2.**
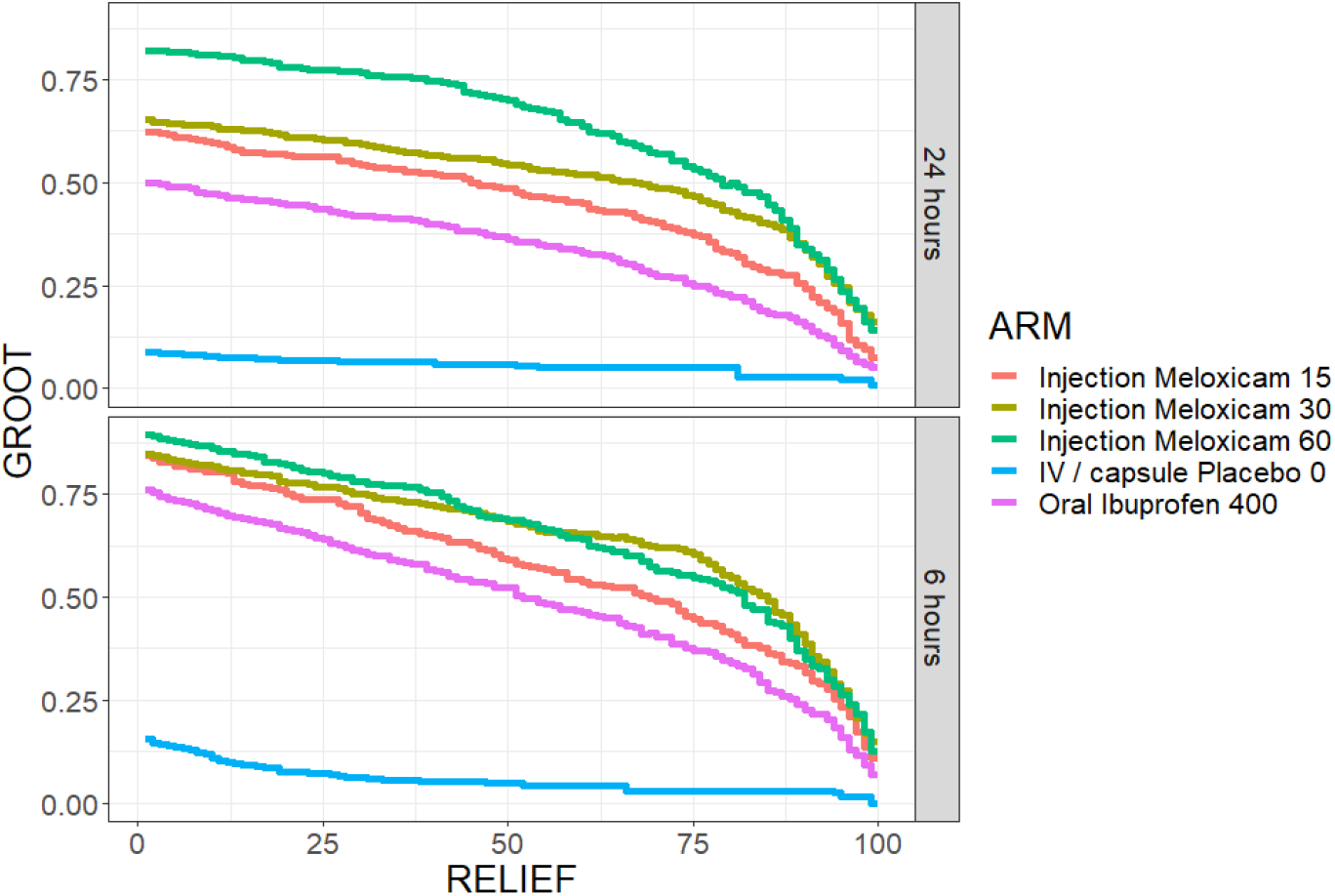
Distribution of GROOT values across different clinically meaningful relief thresholds.

**Figure 3.**
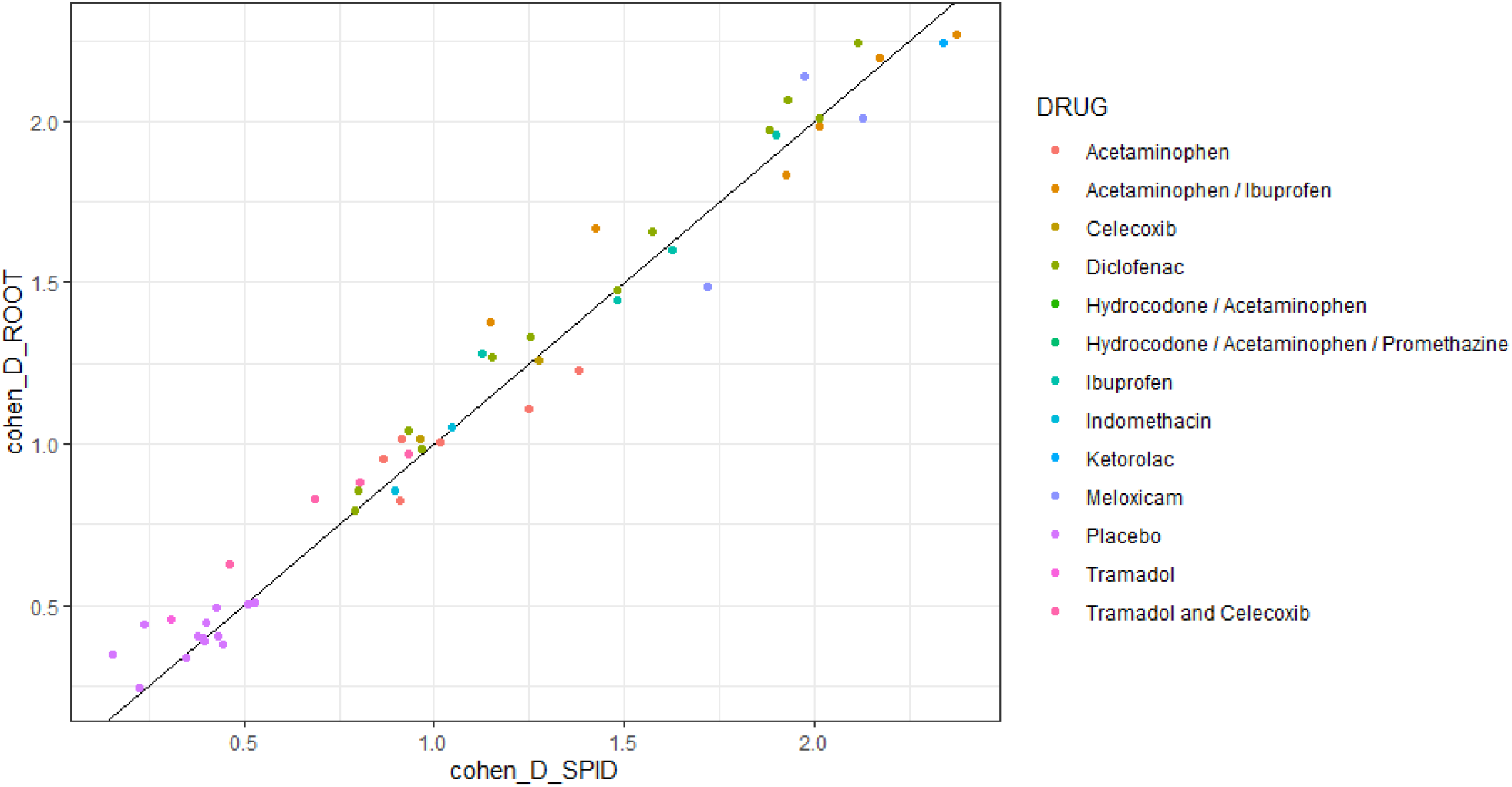
Visual comparison of one-sample Cohen’s D effect size between the SPID and ROOT participant-level outcomes among all study arms. GROOT performs similarly to SPID while offering enhanced interpretability and comparability.

Extending the analysis to 24 hours, the ibuprofen GROOT is now substantially lower than the three IV meloxicam doses due to its shorter duration of action (Figure 1, Table XX). There is also a difference between the 60mg and the 30mg or 15mg doses. A review of the GRO graph (Figure 1) provides the explanation, showing that the 60mg dose maintains the proportion of participants maintaining a response substantially higher for a longer period.

#### 4.3.1 Comparison of GROOT to SPID

To compare the utility of group response as a pain relief index to the standard SPID analysis, we calculated a one-sample Cohen’s D value for all study arms using both outcomes. In all cases, ROOT produced similar Cohen’s D as the SPID. The GROOT summary analysis demonstrated a similar effect size to the SPID (Figure 2, Supplemental Table XX).

### 4.4 Onset & Duration

Onset can be visually assessed in the initial rise of the GRO curves (Figure 1). Time to first achievement of CMR among responders was 43-44 minutes for IV meloxicam and 60 minutes for oral ibuprofen (Table 2). We can also present the distribution of time-to-onset values graphically (Supplemental Figure A2).

The duration of the effect is seen in the decline of the curves after their peak (Figure 1). The median time participants maintained ≥50% relief was 22.5 hours in the 60mg, 15.7 hours in the 30mg, and 14.9 hours in the 15mg IV Meloxicam groups. The meloxicam doses’ duration was substantially longer than the comparator drug ibuprofen, which had a median duration of 7.0 hours (Table 3). The three IV meloxicam treatment groups have different response patterns over time, with the duration of effect correlated with the dose. We also provide a plot of duration across different CMR thresholds in Supplemental Figure A4 for additional information. We observed an artificial multimodality in duration at the end of the study period for most trials, caused by a cluster of patients who did not lose CMR after initial treatment.

**Table 2.** Onset of effect for study treatment as first achievement of a ≥50% reduction in pain. Duration of effect as time spent with ≥50% reduction in pain.

| AMONG RESPONDERS |  | Onset |  |  | Duration |  |  |
| --- | --- | --- | --- | --- | --- | --- | --- |
| Study Arm | Responders | Median Minutes | 25<sup>th</sup> Percentile | 75<sup>th</sup> Percentile | Median Hours | 25<sup>th</sup> Percentile | 75<sup>th</sup> Percentile |
| IV meloxicam 15 mg | 41 / 50 (82%) | **43** | 28 | 88 | **14.9** | 7.0 | 23.1 |
| IV meloxicam 30 mg | 44 / 50 (88%) | **44** | 29 | 59 | **15.7** | 9.9 | 23.1 |
| IV meloxicam 60 mg | 45 / 50 (90%) | **44** | 27 | 88 | **22.5** | 15.3 | 23.5 |
| Oral ibuprofen 400 mg | 38 / 50 (76%) | **60** | 45 | 180 | **20.7** | 5.5 | 20.8 |

## 5 DISCUSSION

Our novel approach to the presentation and analysis of acute pain clinical trial data considers the changing proportion of participants who achieve a prespecified level of clinically meaningful response (CMR) in different treatment arms. The outcomes of group response analysis (GRO, ROOT, GROOT) provide relevant, interpretable information about drug efficacy. The analysis provides a graphical representation and statistical assessment of three clinically and scientifically relevant components of participant response, namely: (1) the speed of onset among responders, (2) the maximal proportion of responders at a clinically meaningful level, and (3) the duration that response. The summary GROOT value provides a measure of overall efficacy as indicated by responder rates over time.

A sum of pain intensity differences (SPID) is a current standard outcome in pain studies, however the SPID is not easily interpreted from a participant response perspective (Wallace, 2023). Furthermore, SPIDs from different studies may have widely different values depending on measurement frequency or study length. Conversely, all GROOTs exist on the same scale: 0 to 1, which facilitates comparative analysis. The calculation of the values over time increases the efficiency matching or exceeding the SPID values.

We demonstrated that GROOT (i.e., mean ROOT), provides a Cohen’s D effect size similar to the SPID. This similarity obviates the frequently voiced concern about the reduction in assay sensitivity from responder analysis, making the GROOT a potentially useful primary outcome for future clinical trials.

Group response analysis’ focus on participant response across time builds on the growing interest in assessing clinical trials using a CMR applied to each participant to define the proportion of participants who respond to therapy rather than group means. GROOT overcomes the primary drawback of a responder analysis: the loss of study power due to dichotomization. The procedure described allows the use of varying methodologies to account for missing data points. In our example, we consider any participant dropping out of the study to be a non-responder after they drop out, and for intermittent missing data, we use the available data before and after the missing point to create a linear interpolation. There are many options that may be flexibly defined, which may allow group response analysis to be applied to a variety of contexts beyond acute pain analgesia.

In the GRO curves for the example of an acute pain clinical trial (figure 1), the GROOT (area under the curves) values for the drug treatment groups are all significantly larger than the 4% for the placebo group (Table XX). In addition, all three doses of IV meloxicam demonstrate faster achievement of ≥50% CID response than oral ibuprofen and differences in the maximum response rate and duration consistent with the circulating drug levels. All these responses are statistically and clinically superior to placebo.

As with any analysis method, the GROOT has limitations. An outcome level must be chosen as an adequate response (i.e., the CMR), but using data over a full range of time values provides greater use of the available data than the endpoint analysis. If the CMR level is not known or there is an interest in looking at a range of cutoff values, the MaxGRO and GROOT can be calculated for all cutoffs and plotted as curves on a graph comparing each arm of the study’s overall cutoff points. (See Supplementary Figures A1 and A3). The use of a single CMR threshold may not accurately reflect the diversity of participant experiences and expectations.

Another consideration in comparing doses or types of medication is the rate of adverse events (AE) that may occur in the selected treatments. A table of treatment-emergent side effects is provided in the publication of this study (Christensen et al., 2018), which showed similar numbers of AEs in all groups, including the placebo group. There were lower rates of nausea and vomiting in the more effective 30mg and 60mg groups, potentially related to less use of the opioid rescue medication. We do not address the issue of risk/ benefit ratios in this paper since the choice of analysis does not alter the process.

However, a more clinically relevant presentation of the efficacy data should facilitate the evaluation.

In conclusion, we have described a simple and understandable method of graphing and analyzing clinical trial data with a summary statistic (GROOT) that estimates the proportion of participants who achieve and maintain a clinically defined response over time. It provides a graphical and tabular representation of the proportion of participants who achieve that response over time, the time to achieving the defined response, and the duration in hours of the response over time. This data analysis provides an efficient estimate of efficacy and provides an easily understood proportion of responder outcome that can directly inform clinicians deciding which medication to use in treating any disease. Due to these favorable properties, we recommend GROOT as a primary or secondary outcome analysis to provide clinically relevant information for the care of patients.

## Data Availability

All data was submitted to the US Food and Drug Administration (FDA) in support of new drug applications, and is held on the FDA's Document Archiving Reporting and Regulatory Tracking System (DAARTS)

## 6 APPENDIX – SUPPLEMENTAL FIGURES

**Figure Supplement A1:**
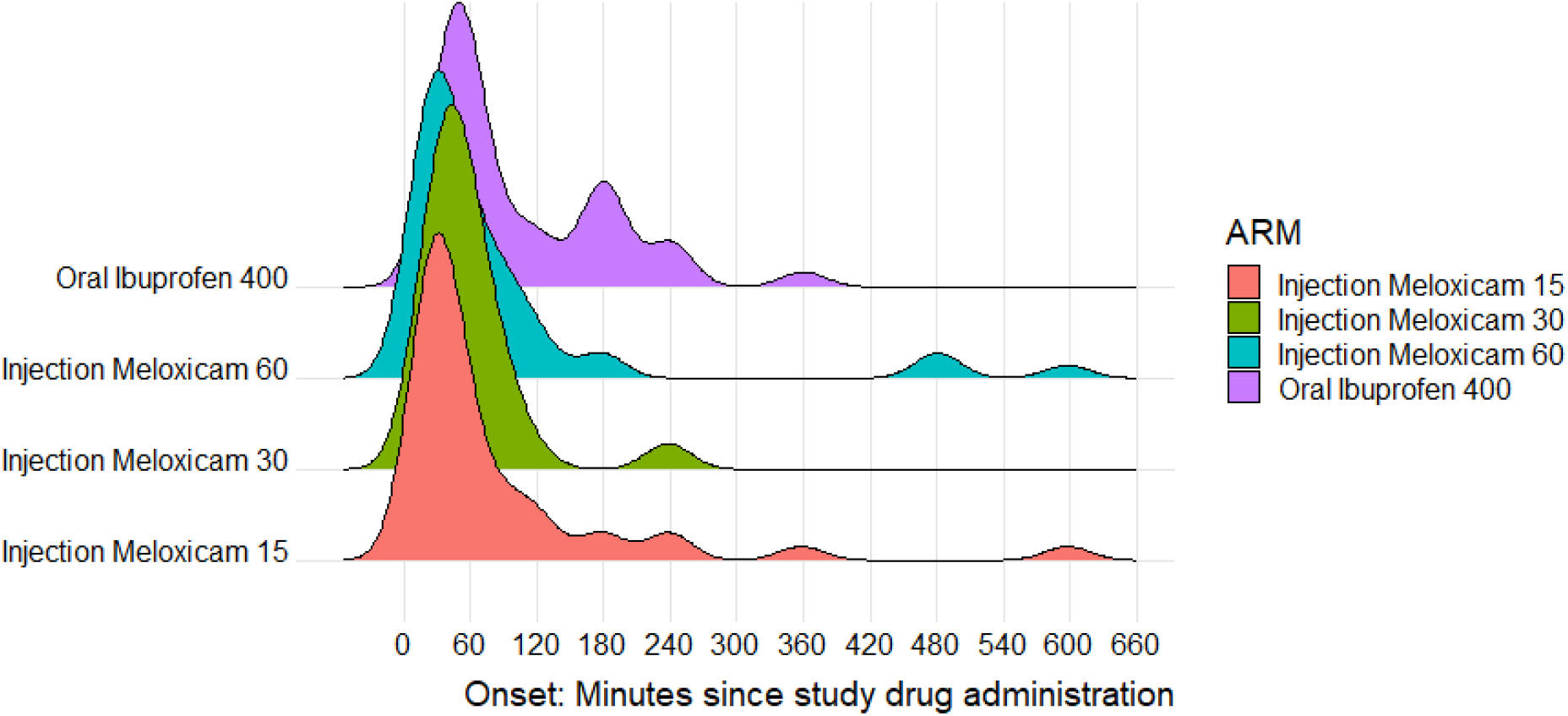
Distribution of time to the first achievement of a clinically important difference in responders.

**Figure Supplement A2:**
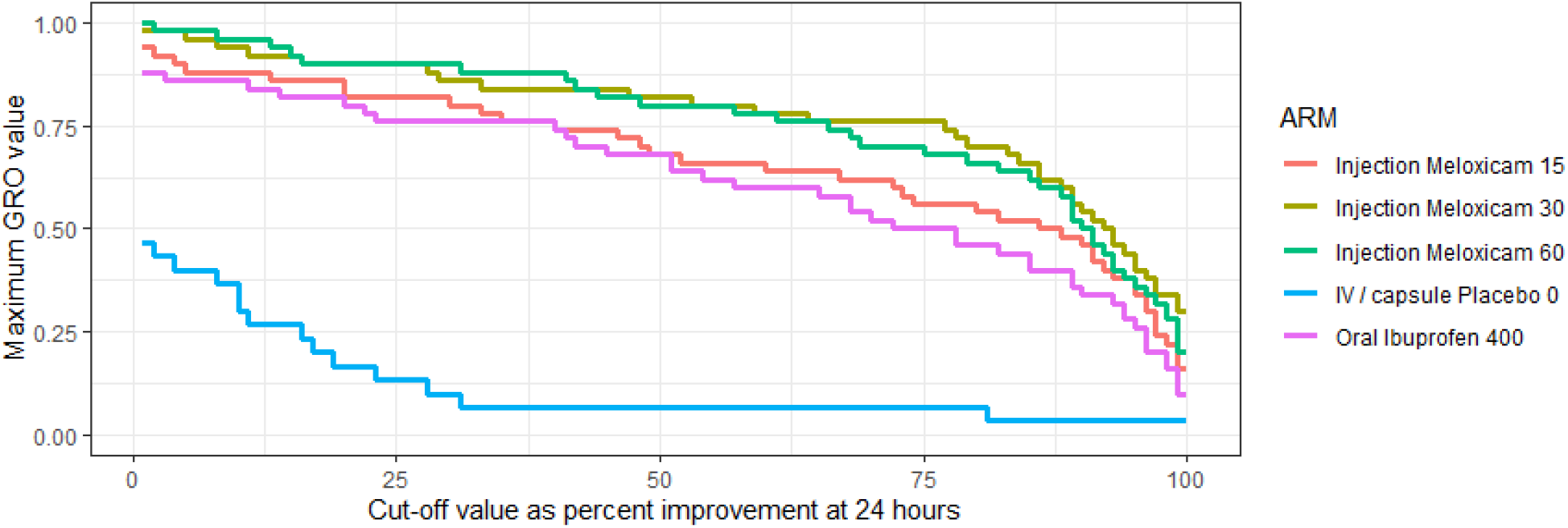
Maximum GRO achieved (MaxGRO) over all cutoff points

**Figure Supplement A3:.**
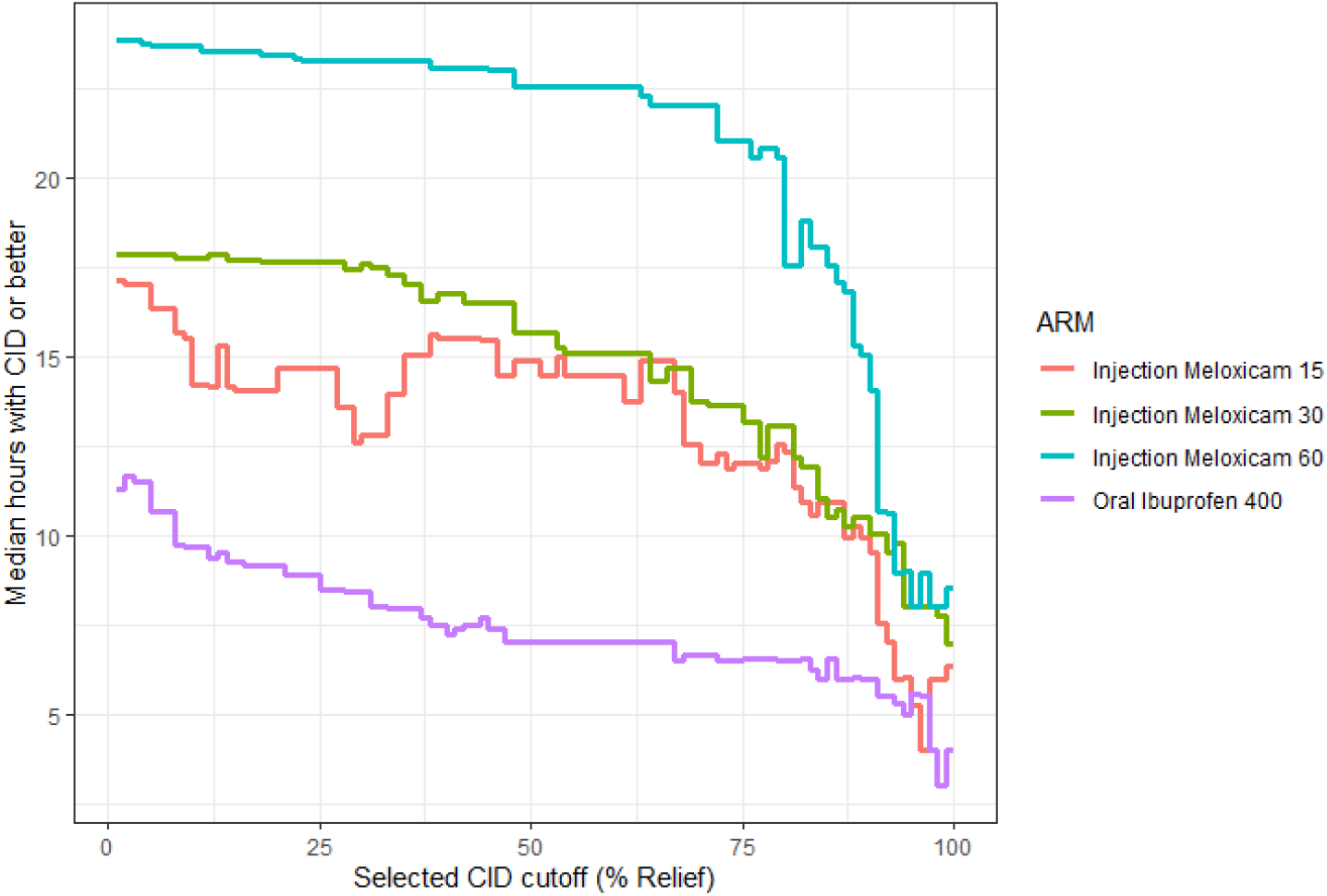
Duration by treatment arm over all cutoff points

## Author contributions

I.D. conceived and developed the foundational end point concept and original analytical framework, and co-developed the FDA database infrastructure that enabled this research. C.J.M. refined the end point methodology for broader applicability. W.B.B. provided statistical expertise and critical methodological review. C.E.A., R.L.B., J.S.G., I.G., J.A.H., and K.N.T. provided clinical expertise and guidance on pain research applications. T.M. contributed to the development of the FDA database infrastructure. J.T.F. provided overall study oversight. All authors provided critical review of earlier drafts and approved the final version.

