## Supplemental materials for "Group Response Analysis: Clinically Interpretable Longitudinal Responder Analysis Methods Developed Using FDA Data"

### Methods

The standard responder analysis examines the difference between the baseline pain level value and another point in time, usually the end point of the study. For this analysis, we use that process to determine the participant's response status from the baseline through each time point in the study, which is graphed as a proportion of responders at every time point (GRO curve) and analyzed as the area under the GRO curve over time (GROOT).

#### The Group Response Outcome (GRO)

To calculate the GRO value for each time point, we determine the response status of each participant at each measured time point and then assess the proportion who are responders at each protocol prespecified time interval over the course of the study. Several components of the analysis need to be specified and disclosed as with any data analysis and are outlined in Table 1 for our example analysis. The two that are particularly important are the choice of the CID to achieve a CMR and the method for handling the interpolation of data across the variable time points logged in the data for each participant's recording of their outcome measure. Given that the times of measurement are often slightly different or missing for some points in the study for each participant, a method for interpolation between time points is important. The specific methods for this estimation are flexible to accommodate for different types of data and are discussed along with other analytic options below.

By graphing the proportion of responders over appropriate time intervals, we are able to visualize: 1) onset (rate of rise in the proportion of responders to the maximum), 2) maximum proportion of responders (maximum height of the GRO curve), and 3) duration of effect (reduction in the proportion of responders over time). The method for the calculation of each is detailed below.

This informative curve (see figure 1) enables a visual comparison of three components of the clinical decision-making process: efficacy, onset, and duration. GRO estimates response rates over time and incorporates these three components to provide an overall assessment of treatment efficacy for the prescribed CMR threshold. Onset is visually represented by the initial rise of the GRO curve. The duration of effect is represented by the curve's plateau.

$$\text{GRO:} \quad g(t \mid \hat{m}) = \frac{1}{n}\sum_{i=1}^{n} \mathbf{1}[\hat{m}_i(t) \in \delta]$$

| | |
|---|---|
| $\mathbf{1}[\cdot]$ | indicator function |
| $\hat{m}_i(t)$ | Efficacy outcome estimate for participant $i$ at time $t$ |
| $n$ | number of participants in the group |
| $\delta$ | set of clinically meaningful response values or categories (see Table 1) |

#### Group Response Outcome over Time (GROOT)

To summarize overall efficacy in a single numeric value, we define the group response outcome over time (GROOT). The GROOT can be calculated as the area under the GRO curve using two methods.

##### *1) GROOT Calculated as the GRO Area as a Proportion of the Total Area.*

The first is by dividing the value under the GRO curve by the total possible area (study length x 100% relief) of the observed time.

$$\text{GROOT:} \quad G(a, b \mid g) = \frac{1}{b-a} \int_a^b g(t)\, \mathrm{d}t$$

| | |
|---|---|
| $g(t)$ | group response outcome (GRO) at time *t* |
| $a$ | time interval start |
| $b$ | time interval end |

Without an easy closed solution, this method requires a bootstrapped resampling of individuals from participant groups an adequate number of times to produce a stable confidence interval.

##### *2) GROOT Calculated as an Average of the Participant-Level Outcomes*

The second is to calculate the period of time each participant meets the CMR condition which we define as the response outcome over time (ROOT) and then average those values over all participants. Alternatively, GROOT can also be derived from a participant-level outcome: the mean proportion of a chosen study period that individual participants qualify as responders. We define this value as the response outcome over time (ROOT) for participant *i* across a time interval specified by (*a, b)*.

$$\text{ROOT:} \quad r_i(a, b \mid \widehat{m}_i) = \frac{1}{b-a} \int_a^b \mathbf{1}[\widehat{m}_i(t) \in \delta]\; dt$$

$$\text{Mean ROOT:} \quad \frac{1}{\mathrm{n}} \sum_{i=1}^{n} r_i(a, b \mid \widehat{m}_i) = \frac{1}{\mathrm{n}} \sum_{i=1}^{n} \frac{1}{b-a} \int_a^b \mathbf{1}[\widehat{m}_i(t) \in \delta]\; dt$$

$$= \frac{1}{b-a} \int_a^b \frac{1}{n} \sum_{i=1}^{n} \mathbf{1}[\widehat{m}_i(t) \in \delta]\, \mathrm{d}t = G(a, b \mid g) \quad \text{GROOT}$$

GROOT's distribution is therefore asymptotically normal, and confidence intervals for GROOT can be generated by any familiar asymptotic method (e.g., Wald or other) which is easier than the bootstrapped resampling procedure of individuals from participant groups for method 1.

#### Calculation of GROOT across all potential CID cut-off points

As with other responder analyses, the GROOT value depends on selecting a CMR cutoff value used to define responders. To generalize the GROOT findings and compare the study arms over a range of cutoff values, we can calculate the GROOT and graph the values over all potential cutoff values.

#### Flexible Options for Analysis

One benefit of our novel methodology is the ease with which it may be applied to diverse analytic contexts. Researchers can make selections based on the needs of their own analysis. Table 1 shows choices we made for the application of group response analysis to the FDA acute pain data, with discussion following.

*Table 1: Choices specific to the group response analysis of the example trials.*

| | |
|---|---|
| **Time Scale** | Hours since study drug administration (continuous variable). |
| **Efficacy Outcome** $m_i(t)$ | Percent change in pain intensity difference between baseline and measurement time reported on the Visual Analog Scale (0-100 VAS) or Numeric Rating Scale (0-10 NRS). |
| **Between-Observations Estimation Method** $\widehat{m}_i(t)$ | Between-point imputation was accomplished using a linear interpolation where $t_{j-1}$ and $t_{j+1}$ are the closest observed efficacy assessments before and after timepoint of interest $t$: $\widehat{m}_i(t) = \begin{cases} m_i(t), & m_i(t) \text{ is observed} \\ m_i(t_{j-1}) + \frac{m_i(t_{j+1}) - m_i(t_{j-1})}{t_{j+1} - t_{j-1}}\{t - t_{j-1}\}, & m_i(t) \text{ is unobserved} \end{cases}$ |
| **Baseline Pain** | Baseline pain is the last observation before study drug administration. |
| **Clincially Important Difference (CID)** $\delta$ | A numeric difference of ≥50% improvement in VAS or NRS using a modified percent change from baseline. Set of CID values $\delta = \{\widehat{m}_i(t) \in \mathbb{R} \mid \widehat{m}_i(t) \leq -0.50\}$ Where $\widehat{m}_i(t)$ is the estimated VAS/NRS estimate for patient *i* at time *t.* For this example, negative values indicate improvement from baseline. |
| **Missing Data Policy** | Participants who withdraw or take rescue are treated as non-responders for all time points after the event (baseline observation carry forward - BOCF). |

#### Time Scale and Baseline

GRO estimates the changing level of response over time in the clinical trial, from the defined baseline at time zero, to a later study timepoint. It can use any reasonable time scale. For acute pain analgesia, the initial dose of medication administration is an obvious choice for baseline time zero.

#### Efficacy Outcome and Acceptable CMR Thresholds

These methods require the *a priori* definition of a clinically meaningful response (CMR). This selection is then applied to individual participant data to dichotomously categorize subjects as a responder or not at each timepoint during the study, using a suitable unobserved timepoint estimation method. ROOT, GRO & GROOT can be calculated for multiple defined outcome levels or compared across the full range of values to explore a range of results. (Supplemental Figure A1).

Depending on the specific application, outcomes may be raw measurements or derived values such as change from baseline. Percent changes from baseline are common among responder analyses in pain studies. {#REF} Still, researchers must carefully consider the ceiling and floor effect of specific scales and consider using modified percent change outcomes to address the issue that percent changes up and down do not correspond to the same absolute difference.

#### Missing Data

Participants can withdraw from trials at any time, so the analysis must account for missing data. We chose to treat patients who took rescue, withdrew, or whose observations otherwise ended early, as non-responders after the time of their withdrawal or missingness; as in an Intent-to-Treat (ITT) analysis. However, any appropriate missing data method can be adapted. Although it may be that some of those dropping out for side-effects may have experienced successful treatment, the conservative approach is to assume they were non-responders. This prevents bias toward a larger responder rate within study arms. This also reflects the clinical situation where patients who experience significant side effects would likely not want to continue with a treatment even if it was effective.

#### CMR Status Estimation Between Observations

A GRO that changes only when a participant is actually observed is unlikely to accurately reflect truth, since responder status change likely occurs in the period between measurements. For continuous outcomes, a linear interpolation of values over time is simple and often appropriate, but any reasonable method may be chosen to estimate these values. Categorical outcomes cannot employ the same estimation methods, at least not without establishing a linear relationship between the categories or employing more sophisticated methods.

### Analysis of Onset & Duration

Achievement of CMR can be useful in the analysis of the onset and duration of treatment efficacy. However, simply reporting summary statistics for these values may not provide answers that are clinically relevant. Some participants do not respond to treatment, and for them the concepts of onset and duration cannot be meaningfully applied. One analytical option is to restrict analysis of onset and duration to responders only. In this case, the analyses must be considered observational in nature.

Under this framework, onset is summarized as median time to first achievement of CMR. Duration can be summarized using the time that individuals spend as responders. These definitions are consistent with ubiquitous clinical questions: (1) how long after treatment can the patient expect response?; (2) how long can a responder expect the response to last if they achieve a response? In addition to the median time value, another useful presentation is a cumulative function of the proportion of responders with durations at or above the time represented on the x-axis (Supplemental Figure XX). This provides data to allow clinicians to make an informed decision about the timing of redosing based on the likely time of return of pain in the population, while balancing the increased risk of experiencing adverse events with higher drug exposure.

Observational cohorts based on the post-randomization factor of responder status includes risk of confounding. Note also that duration can only be meaningfully measured if the study duration is longer than the effect of the treatment. In many pain models, it is reasonable to expect a small but significant number of participants will retain CMR for the entire study period. This may constitute informative censoring of duration times, which researchers should consider as they select methods for the analysis of the onset & duration data. A Cure model {#REF Cure model} for onset or duration may be a reasonable approach to preserve randomization, handle censoring, and use data from all participants but is beyond the scope of this paper.
